# Proteomic Age Acceleration in Multiple Sclerosis Precedes Symptom Onset and Associates with Severity

**DOI:** 10.64898/2026.04.13.26350634

**Authors:** Fatemeh Siavoshi, Julián Candia, Dimitrios C. Ladakis, Blake E. Dewey, Angeliki Filippatou, Matthew D. Smith, Elias S. Sotirchos, Shiv Saidha, Jerry L. Prince, Ahmed Abdelhak, Ellen M. Mowry, Peter A. Calabresi, Keenan A. Walker, Kathryn C Fitzgerald, Pavan Bhargava

## Abstract

Biological aging is accelerated in people with multiple sclerosis, but whether such acceleration occurs during the pre-symptomatic phase or varies by organ system is understudied.

We analyzed two independent proteomics datasets profiled using distinct platforms: the Johns Hopkins cohort profiled using the SomaScan platform (348 multiple sclerosis/49 age-matched controls) and the Department of Defense cohort profiled using the Olink platform (134 multiple sclerosis/79 age-matched controls), including 117 pre-symptomatic samples from people with multiple sclerosis (median lead time: 4.0 years), to estimate systemic and organ-specific proteomic age gaps using established clocks in pre-symptomatic and symptomatic phases, and assess their associations with severity.

In the Johns Hopkins cohort, people with multiple sclerosis demonstrated acceleration of systemic (β=2.2, 95% CI 1.2–3.2, *P*<0.001, FDR<0.001), brain (β=1.7, 95% CI 0.6–2.7, *P*=0.003, FDR=0.01), muscle (β=2.5, 95% CI 1.3–3.7, *P*<0.001, FDR<0.001), and immune age (β=1.8, 95% CI 0.6–2.9, *P*=0.003, FDR=0.01), with findings reproduced in the Department of Defense cohort for systemic (β=0.7, 95% CI 0.0–1.4, *P*=0.04, FDR=0.34) and brain age (3.2 years, 95% CI 2.1–4.3, *P*<0.001, FDR<0.001). Proteomic age acceleration was evident prior to symptom onset [systemic: (β=1.0, 95% CI 0.4–1.7, *P*=0.002, FDR=0.02); brain: (β=2.4, 95% CI 1.2–3.7, *P*<0.001, FDR=0.002)], whereas no immune age acceleration was detected before or after onset. Higher systemic age gap was associated with greater global Age-Related Multiple Sclerosis Severity Score (β=0.14, 95% CI 0.05–0.24, *P*=0.005, FDR=0.03) and slower walking speed (β=0.02, 95% CI 0.01–0.03, *P*=0.006, FDR=0.04), while higher muscle age gap was associated with greater global Age-Related Multiple Sclerosis Severity Score (β=0.17, 95% CI 0.10–0.24, *P*<0.001, FDR<0.001), poorer manual dexterity (β=0.28, 95% CI 0.04–0.52, *P*=0.03, FDR=0.30), slower walking speed (β=0.02, 95% CI 0.01–0.03, *P*=0.002, FDR=0.02), lower peripapillary retinal nerve fiber layer (β= −0.26, 95% CI −0.41 to −0.10, *P*=0.001, FDR=0.02) and ganglion cell-inner plexiform layer thicknesses (β= −0.35; 95% CI −0.65 to −0.05; *P*=0.02, FDR=0.30). Higher brain age gap was associated with several imaging measures, including lower whole-brain (β= −0.002, 95% CI −0.003 to −0.001, *P*=0.002, FDR=0.02), and lower peripapillary retinal nerve fiber layer thickness (β= −0.21, 95% CI −0.39 to −0.03, *P*=0.02, FDR=0.10).

Proteomic age acceleration in multiple sclerosis is detectable years before symptom onset and distinct organ-specific aging signatures are associated with disease severity. Proteomic aging may provide a biologically informative marker of early disease processes and a clinically relevant readout of disease heterogeneity.

## Introduction

Multiple sclerosis is a chronic inflammatory and neurodegenerative disease of the central nervous system with highly variable onset and clinical course. Chronological age is one of the strongest determinants of phenotype, influencing relapse frequency, response to therapy, and the transition to progressive disability, independent of disease duration. Yet individuals of the same chronological age can follow markedly different trajectories, suggesting that inter-individual differences in biological aging at the molecular level may contribute to heterogeneity in multiple sclerosis outcomes and more accurately capture vulnerability to tissue damage or capacity for repair^1^.

Recent proteomic aging models (“proteomic clocks”), developed from large population cohorts, demonstrate robust prediction of chronological age and strong associations with age-related morbidity and mortality, underscoring their utility as integrative biomarkers of biological aging. By quantifying age-related changes across hundreds to thousands of circulating proteins, these models may capture downstream effects of immune, metabolic, and vascular processes that are highly relevant to multiple sclerosis pathophysiology^2,3^. Growing evidence indicates that biological aging processes are accelerated in multiple sclerosis, including disease-modifying therapy–naïve adults and pediatric-onset cases. Studies have reported shorter telomere length, epigenetic age acceleration, metabolomic aging signatures, and other age-related molecular changes in people with multiple sclerosis, which in turn have been linked to greater disability and brain atrophy^4–7^. However, it remains unclear when this acceleration emerges and whether it reflects a consequence of established disease pathology, or an earlier preclinical process that precedes symptom onset.

The pre-symptomatic phase (prodrome) of multiple sclerosis may offer a unique opportunity to examine the temporal relationship between biological aging and disease onset, although biological aging during this period remains largely unexplored. To address this gap, we applied established proteomic aging models to two independent serum proteomics cohorts, spanning distinct proteomic platforms, one of which leveraged pre-symptomatic samples from individuals who later developed multiple sclerosis. By comparing proteomic age between people with multiple sclerosis and age-matched healthy individuals across these cohorts, we sought to determine (1) whether systemic and organ-specific proteomic age acceleration is present in multiple sclerosis, (2) whether this acceleration is detectable before the first clinical manifestation of disease, and (3) whether proteomic age gaps relate to clinical and imaging measures of disease severity.

## Materials and methods

### Study participants

We analyzed serum proteomic datasets from two independent cohorts; an overview of the design and analysis is provided in Figure 1. The first cohort included individuals with multiple sclerosis at various disease stages and age-matched healthy controls recruited through the Johns Hopkins Multiple Sclerosis Precision Medicine Center of Excellence (JHU). Participants with multiple sclerosis included individuals with relapsing–remitting and progressive multiple sclerosis, who were receiving disease-modifying therapies (DMTs) of varying efficacy. DMTs at the time of blood draw were categorized as high-efficacy (natalizumab, ocrelizumab, rituximab), moderate-efficacy (fingolimod, dimethyl fumarate), or mild-efficacy (glatiramer acetate, interferon-β, terfluoronamide, mycophenolate mofetil). Relapsing-remitting and progressive multiple sclerosis diagnoses were assigned according to the 2017 McDonald criteria^8^. Blood samples were collected using standardized protocols and stored at −80 °C until analysis.

**Figure 1.**
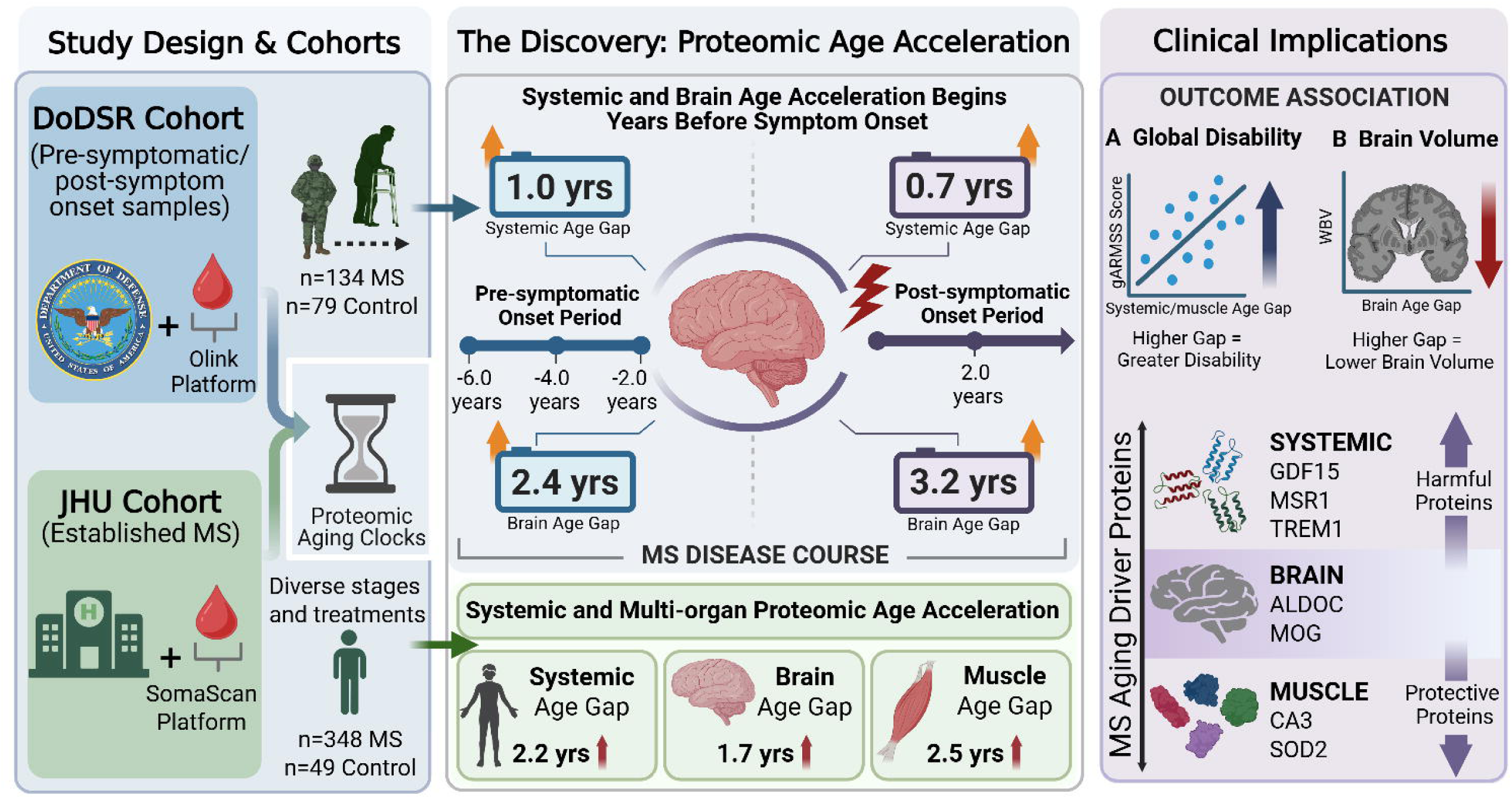
Overview of the study design, cohorts, proteomic platforms, and principal findings.

We analyzed a publicly available serum proteomic dataset derived from the Department of Defense Serum Repository (DoDSR) pre-symptomatic multiple sclerosis cohort, which has been described previously^9^. This cohort was established to characterize biological changes preceding multiple sclerosis onset within the U.S. military population. Briefly, the parent cohort includes 250 individuals with multiple sclerosis identified from the Gulf War Era MS (GWEMS) study, a population-based cohort of incident multiple sclerosis cases occurring during active-duty service between 1990 and 2007^10^. Multiple sclerosis diagnoses were confirmed according to the 2017 McDonald criteria^8^. For each multiple sclerosis case, two longitudinal serum samples were selected: the earliest available pre-symptomatic sample (median lead time −4.0 years [range: -14 to -1 years]) and a second sample collected shortly after symptom onset (median lead time 2.0 years [range: 1 to 6 years]). Each case was matched to a healthy control donor based on age, sex, race/ethnicity, military branch, and dates of serum collection. The de-identified dataset used in the present study was obtained from the Dryad digital repository^11^.

All participants provided written consent in accordance with the Declaration of Helsinki, and the study was approved by the Johns Hopkins Institutional Review Board.

### Proteomics analysis

Proteins from JHU serum samples were measured using the SomaScan v4.1 platform (7,288 SOMAmer reagents), as described previously^12–14^. Using 102 blind duplicates, proteins with intra-assay coefficients of variation (CV) >50% were excluded (n=20). Samples from participants that did not pass SomaScan quality control (QC) criteria were also excluded (n=7). Values were log_2_ transformed, and those beyond 5 standard deviations (SDs) were winsorized.

Serum samples from the DoDSR cohort were analyzed using the Olink Explore HT platform. Following centrifugation at 10,000 g for 5 minutes, 50 µl aliquots were transferred to 96-well plates along with internal, plate, and negative controls. Protein levels were quantified using the proximity extension assay from the Explore HT panel (∼5,400 proteins), and sequencing was carried out on an Illumina NovaSeqX instrument. Normalized protein expression (NPX) values were generated after adjustment to five inter-plate controls. Measurement precision was assessed from three control samples on each plate, and sample quality was monitored using incubation, extension, and amplification controls included in every well. Assays in which the median NPX of the negative controls deviated by more than five SDs from predefined thresholds were flagged for quality concerns. Potential outliers were identified using principal component analysis, applying a ±5 SD threshold around the aggregated mean NPX distribution. Any sample or assay failing these quality criteria was removed before downstream analysis^15^.

### Clinical and imaging outcome measures

Disease severity in the JHU cohort was assessed using the Global Age-Related Multiple Sclerosis Severity Score (gARMSS)^16^, quantitative functional performance testing, retinal imaging, and structural brain MRI. gARMSS, retinal imaging, and MRI measures were included if obtained within 6 months of proteomic blood sampling, and quantitative functional assessments were included if performed within 1 month.

Quantitative functional performance was measured using an iPad-based assessment of upper extremity function, ambulation, and information processing speed, including the Manual Dexterity Test, Walking Speed Test, and Processing Speed Test respectively^17–19^.

Retinal imaging was performed using spectral-domain Cirrus HD-OCT instruments (models 4000, 5000, and 6000; Carl Zeiss Meditec, Dublin, California). Optic disc and macular scan protocols were performed as previously described, and quality review was in accordance with OSCAR-IB quality control criteria^20^. To reduce variability related to scanner model and software acquisition version, OCT measures were harmonized across devices using the longitudinal ComBat^21^. Retinal layer segmentation was generated using a previously validated automated pipeline^22^.The primary retinal outcomes were ganglion cell–inner plexiform layer (GCIPL) and peripapillary retinal nerve fiber layer (pRNFL) thicknesses. Participants were excluded from OCT analyses if they had medical or ophthalmologic conditions that could interfere with OCT testing or results, including diabetes mellitus, uncontrolled hypertension, glaucoma, prior ocular trauma or surgery, high refractive errors greater than ±6 diopters, or other major neurologic or ophthalmologic disorders.

Brain MRI was acquired on multiple research and clinical scanners using a standardized whole-brain protocols. The core sequences included three-dimensional sagittal magnetization-prepared rapid gradient echo (MPRAGE; 1 mm isotropic), three-dimensional sagittal fluid-attenuated inversion recovery (FLAIR; 1 mm isotropic), and multislice T2-weighted dual-echo turbo spin echo imaging (in-plane resolution 0.8–1 mm; slice thickness 3–5 mm). Our MRI processing workflow was designed for use in large, multi-center multiple sclerosis clinical trial that relies on clinically acquired imaging data. By applying advanced methods such as super-resolution and image harmonization, the pipeline enables consistent analysis across scanners, improving statistical power and allowing more accurate modeling of longitudinal changes by incorporating scans obtained over time from different imaging platforms. Images were harmonized before downstream analysis using the Harmonization with Attention-based Contrast, Anatomy, and Artifact Awareness (HACA3) framework^23–25^. Regional brain volumetric measures were then derived using an automated segmentation pipeline described previously^26^. White matter lesions were segmented with a deep learning approach based on the Self-Ensembled Lesion Fusion (SELF) framework^27^. Intracranial volume (ICV) was estimated using a deep neural network trained to identify the inner skull boundary^28^. All regional volumes were normalized to ICV to account for between-subject differences in head size. For bilateral regions, left and right normalized volumes were summed. Composite measures, including cortical gray matter, subcortical gray matter, and whole-brain volume fractions, were calculated by combining their respective component regions.

### Statistical analysis

#### Proteomic age calculation

Proteomics data from these cohorts were used to calculate proteomic age using previously published proteomic aging models developed based on the SomaScan and Olink platforms^2,29^. For the SomaScan dataset, we applied a proteomic aging clock derived from a large-scale human plasma proteomic profiling study trained on 4,778 plasma proteins across 5,676 participants. The Olink-based aging clock was developed using 2,916 plasma proteins quantified in 44,498 individuals from the UK Biobank. Briefly, organ-specific proteomic aging models were constructed by identifying organ-enriched plasma proteins using human tissue bulk RNA sequencing data from the Genotype-Tissue Expression (GTEx) atlas^30^. Genes were considered organ-enriched if their expression in one organ exceeded that in all other organs by at least fourfold. Because the immune system is not represented as a single GTEx tissue, it was defined based on gene expression patterns observed in blood and spleen. These organ-enriched genes were then mapped to plasma proteins measured by SomaScan or Olink, allowing assignment of proteins to specific organs, while proteins without clear organ enrichment were categorized as “organismal.” These clocks enabled estimation of systemic proteomic age as well as organ-specific biological ages for the brain, heart, liver, kidney, lung, muscle, pancreas, adipose, intestine, artery, organismal, and immune systems.

#### Descriptive and analytical models

Descriptive analyses were first performed to summarize demographic and clinical characteristics of each cohort. Continuous variables were presented as mean ± SD or median, interquartile ranges (IQR) based on distributional properties, and categorical variables were reported as counts and percentages. Within each cohort and for each aging clock, Pearson correlation coefficients were calculated to quantify the relationship between chronological age and predicted proteomic age. Proteomic age gap was derived according to the published framework. For the SomaScan data, age gaps were computed using the OrganAge Python package. For the Olink dataset, proteomic age gap was defined as the residual from linear regression of predicted proteomic age on chronological age (Figure 2A). Proteomic age gap served as the primary outcome for downstream analyses.

**Figure 2.**
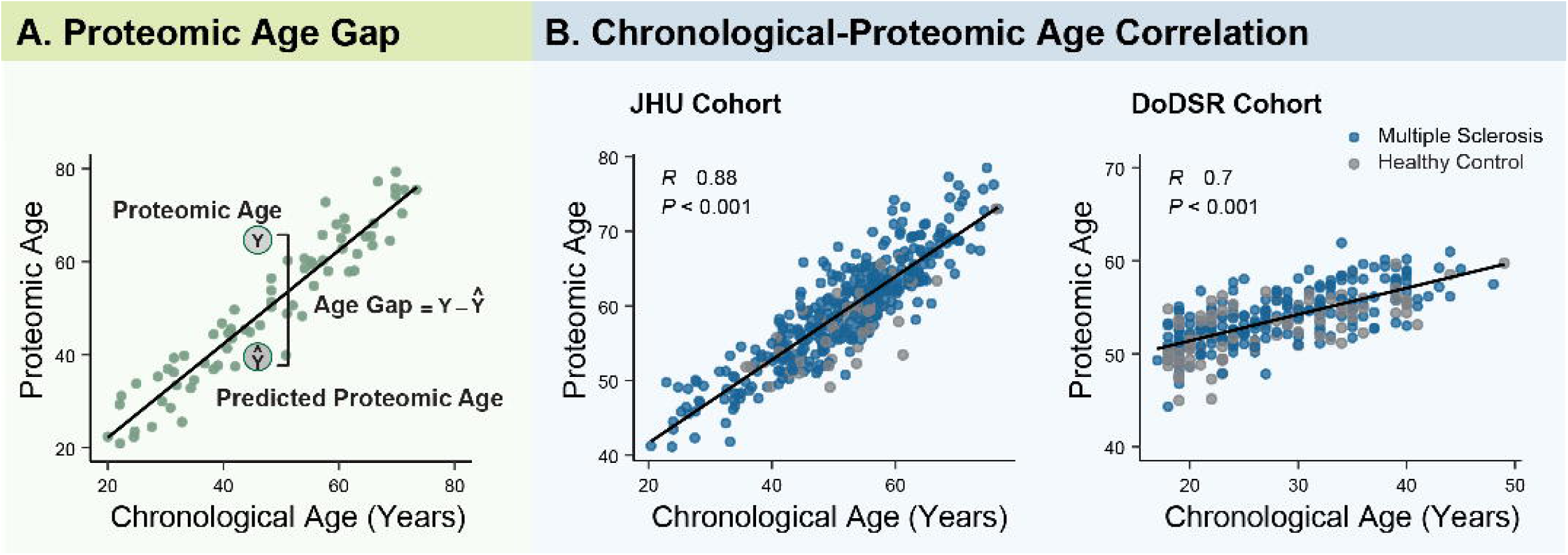
Calculation of proteomic age gap and correlation between chronological age and proteomic age across proteomic aging clocks. **(A)** Schematic illustrating how proteomic age acceleration (proteomic age gap) was calculated as the difference between predicted proteomic age and chronological age, such that positive values indicate accelerated biological aging relative to chronological age. **(B)** Scatterplots showing the relationship between chronological age and predicted proteomic age across the proteomic aging clocks in the JHU and DoDSR cohorts. Each point represents one sample; Pearson correlation coefficients indicate agreement between chronological age and predicted proteomic age.

To evaluate whether individuals with multiple sclerosis exhibit accelerated proteomic aging, multivariable linear regression models were fitted with proteomic age gap as the dependent variable and multiple sclerosis status as the primary independent variable. Models were adjusted for chronological age, sex, and race in the JHU cohort, and for chronological age and sex in the DoDSR cohort, as these were the available demographics. Separate models were fitted for each platform-derived aging clock within each cohort. To assess whether proteomic age acceleration was detectable prior to clinical symptom onset, these analyses were repeated using the earliest available pre-symptomatic samples. Within the DoDSR cohort, exploratory analysis restricted to individuals who developed multiple sclerosis was conducted to examine whether proximity to symptom onset was associated with proteomic age acceleration. Multivariable linear regression models were fitted with proteomic age gap as the outcome and time (in years) to first clinical symptom onset as the primary predictor, adjusting for chronological age at sampling and sex. We also conducted subgroup analyses restricted to individuals with multiple sclerosis. Subgroups of interest included sex, race, multiple sclerosis disease course (relapsing-remitting vs progressive), and DMT class (low-, moderate-, and high-efficacy DMT). In the JHU cohort, associations were evaluated using linear regression models adjusted for relevant covariates. Sex-associated differences were assessed by including sex as a predictor in models adjusted for age, race, and multiple sclerosis subtype; race-associated differences were assessed in models adjusted for age, sex, and multiple sclerosis subtype. Multiple sclerosis subtype analyses were adjusted for age, sex, and race, and DMT analyses were adjusted for age, sex, race, and multiple sclerosis subtype. In the DoDSR cohort, sex-associated differences were assessed using linear regression models adjusted for age and sex.

To identify proteins driving the observed age acceleration in multiple sclerosis subjects relative to controls, the contribution of each protein was calculated for every participant as the product of its normalized abundance and the corresponding model coefficient (i.e., the trained weight from the published aging model), representing its weighted contribution to predicted biological age. In the JHU cohort, multivariable linear regression models were fitted separately for each protein, with protein contribution as the dependent variable and multiple sclerosis status as the primary independent variable, adjusting for chronological age, sex, and race. Proteins with a *P* < 0.05 were considered candidate drivers of accelerated aging within their respective organ-specific clocks.

Associations between age gap values and severity outcome measures were examined using multivariable linear regression models adjusted for chronological age, sex, and race.

All statistical analyses were performed using R version 4.4.1 and Python 3.10.19. False discovery rate (FDR) correction using the Benjamini–Hochberg method was applied within each set of analyses. Due to the exploratory nature of these analyses, statistical significance was defined as nominal P < 0.05, with FDR-adjusted P values also reported.

## Results

### Demographics and clinical characteristics

The JHU cohort included 348 individuals with multiple sclerosis and 49 healthy controls (Table 1). The mean age at sample collection was 51.6 ± 11.2 years among individuals with multiple sclerosis and 51.6 ± 7.9 years among healthy controls. The multiple sclerosis group consisted of 247 females (71.0%) and 101 males (29.0%), and the healthy control group included 35 females (71.4%) and 14 males (28.6%). Among individuals with multiple sclerosis, 278 (79.9%) were White and 61 (17.5%) were Black, and 39 (79.6%) healthy controls were White and 9 (18.4%) were Black. Within the multiple sclerosis group, 247 individuals (71.0%) had relapsing–remitting multiple sclerosis, and 101 individuals (29.0%) had progressive multiple sclerosis.

**Table 1.**
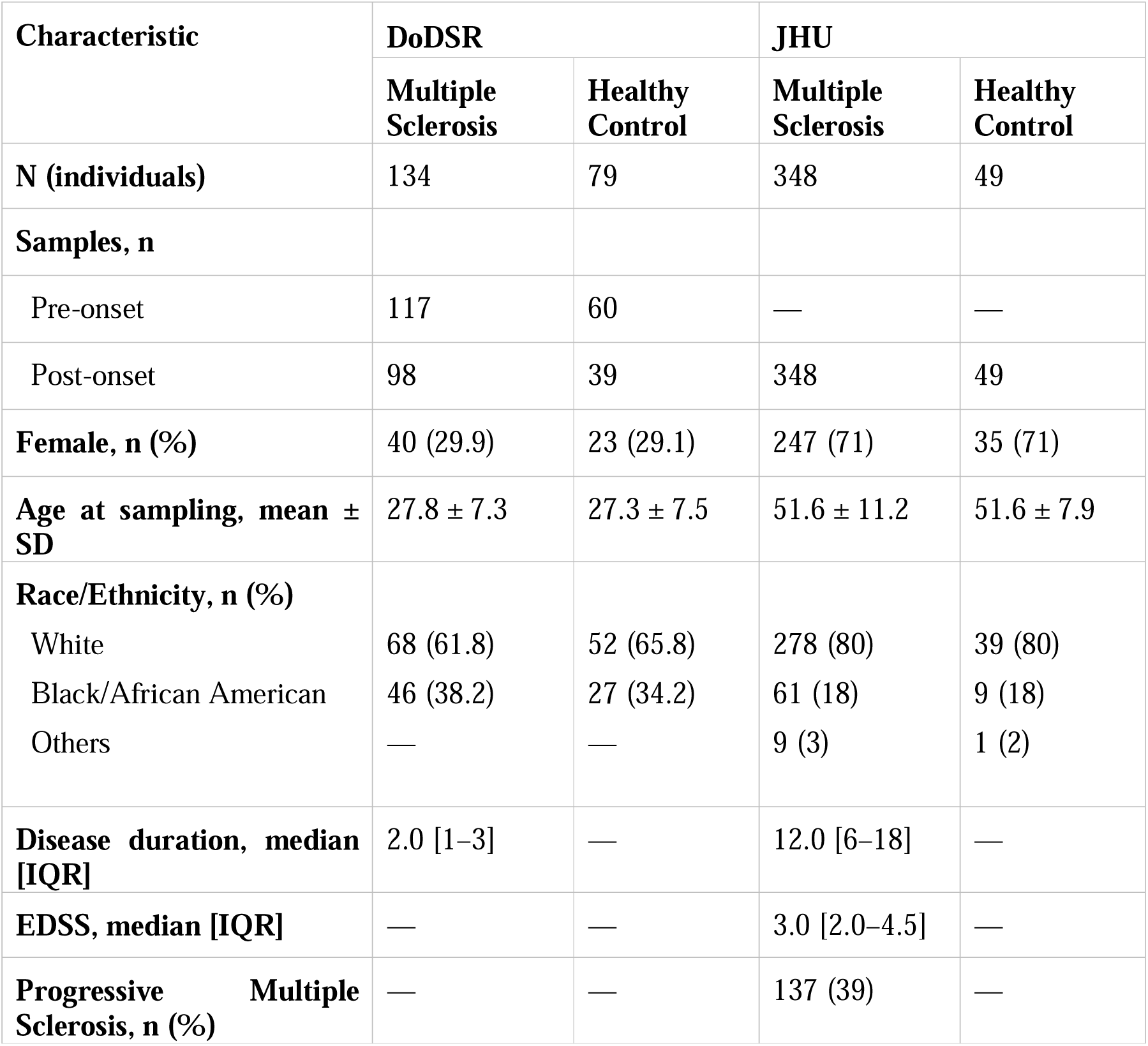
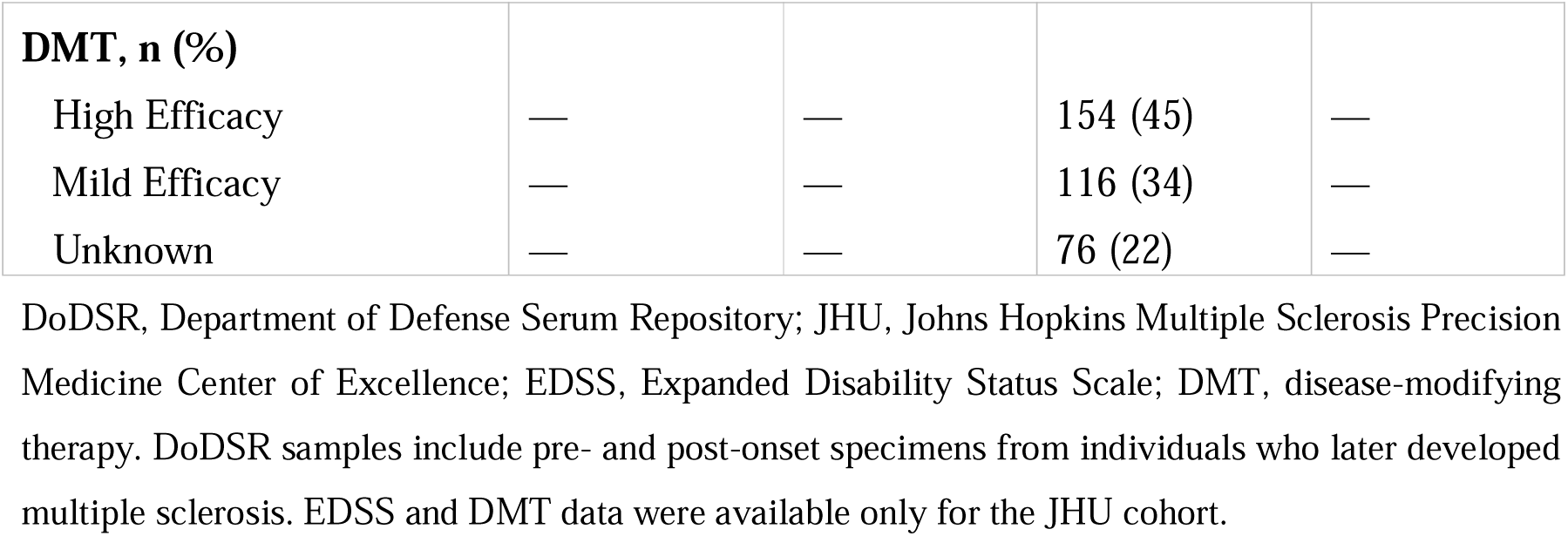
Demographic and clinical characteristics of the study cohorts.

The DoDSR cohort comprised 134 individuals with multiple sclerosis (Table 1), contributing 117 pre-clinical onset samples (median lead time −4.0 years; IQR −6.0 to −2.0), and 98 post-onset samples (median lead time 2.0 years; IQR 1.0 to 3.0). The healthy control group included 79 age-matched individuals, from whom 99 total samples were available. Among individuals with multiple sclerosis, the mean age at sample collection was 24.7 ± 6.2 years for pre-clinical onset samples and 31.4 ± 6.9 years for post-onset samples. Healthy controls were age-matched to the multiple sclerosis group at each sampling timepoint, with mean ages of 24.8 ± 6.6 and 31.1 ± 7.2 years, respectively. The multiple sclerosis group included 40 females (29.9%), and the healthy control group included 23 females (29.1%). Among healthy controls, 27 individuals (34.2%) were Black and 52 (65.8%) were White. Among individuals with multiple sclerosis, 46 (38.2%) were Black and 68 (61.8%) were White. The detailed characteristics of the included participants for the cohorts are shown in Table 1.

### Proteomic age

There was a strong correlation between chronological and systemic proteomic age across both aging clocks (SomaScan clock: r = 0.88, *P* < 0.001; Olink clock: r = 0.70, *P* < 0.001; Pearson correlation) (Figure 2B).

In the JHU cohort (n = 348 multiple sclerosis; n = 49 controls), analysis using the SomaScan-derived proteomic aging clock demonstrated that people with multiple sclerosis exhibited a higher systemic age gap than controls by 2.2 years (95% CI 1.2 to 3.2; *P* < 0.001; FDR < 0.001). Brain age gap was also higher in people with multiple sclerosis by 1.7 years (95% CI 0.6 to 2.7; *P* = 0.003; FDR = 0.01), and immune age gap was increased by 1.8 years relative to controls (95% CI 0.6 to 2.9; *P* = 0.003; FDR = 0.01), independent of chronological age, sex, and race. Muscle age gap was similarly higher in people with multiple sclerosis by 2.5 years (95% CI 1.3 to 3.7; *P* < 0.001; FDR < 0.001). Comparable differences were observed across several organ-specific clocks, including organismal (β = 2.2 years; 95% CI 1.2 to 3.2; *P* < 0.001; FDR < 0.001), pancreas (β = 1.6 years; 95% CI 0.6 to 2.7; *P* = 0.002; FDR = 0.01), liver (β = 1.6 years; 95% CI 0.4 to 2.8; *P* = 0.007; FDR = 0.02), adipose tissue (β = 1.5 years; 95% CI 0.5 to 2.6; *P* = 0.005; FDR = 0.02), intestine (β = 1.4 years; 95% CI 0.4 to 2.4; *P* = 0.008; FDR = 0.02), heart (β = 1.1 years; 95% CI 0.2 to 2.1; *P* = 0.02; FDR = 0.04), and artery (β = 1.1 years; 95% CI 0.0 to 2.3; *P* = 0.05; FDR = 0.08). No significant differences were observed for kidney age gap (β = 0.3 years; 95% CI −0.3 to 0.9; *P* = 0.31; FDR = 0.39) or lung age gap (β = 0.2 years; 95% CI −0.6 to 1.1; *P* = 0.60; FDR = 0.62) (Figure 3).

**Figure 3.**
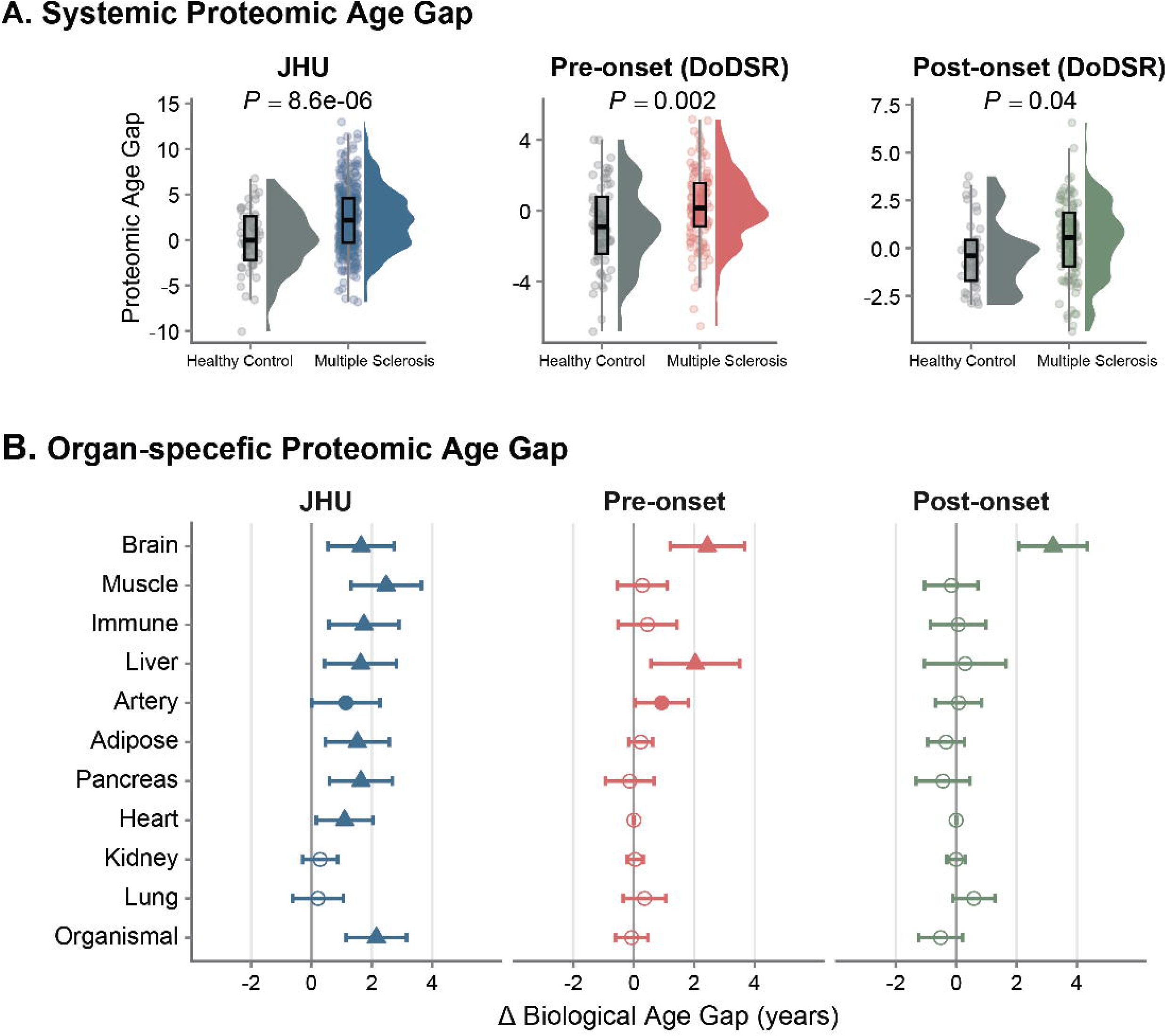
Systemic and organ-specific proteomic age gaps in established, pre-onset, and post-onset multiple sclerosis. (A) Systemic proteomic age gap in people with multiple sclerosis and healthy controls across three comparisons: established multiple sclerosis in the Johns Hopkins Multiple Sclerosis Center (JHU) cohort, pre-onset multiple sclerosis in the Department of Defense Serum Repository (DoDSR) cohort, and post-onset multiple sclerosis in the DoDSR cohort. Positive values indicate accelerated proteomic aging relative to chronological age. (B) Organ-specific proteomic age gap differences between people with multiple sclerosis and healthy controls across the same three disease stages: established multiple sclerosis, pre-onset multiple sclerosis, and post-onset multiple sclerosis. Values shown represent the estimated difference in biological age gap (years) between multiple sclerosis and healthy control for each organ-specific clock. Filled circles indicate nominal significance (P < 0.05), and triangles indicate associations that remain significant after false discovery rate (FDR) correction (FDR-adjusted P < 0.05).

Comparable findings, particularly for systemic and brain proteomic aging, were observed in the DoDSR cohort using Olink-based aging models applied to post-onset samples (n = 98 multiple sclerosis; n = 39 controls). Systemic proteomic age gap was higher in individuals with multiple sclerosis by 0.7 years (95% CI 0.0 to 1.4; *P* = 0.04; FDR = 0.34), while brain proteomic age gap was substantially higher by 3.2 years (95% CI 2.1 to 4.3; *P* < 0.001; FDR < 0.001). Differences in proteomic aging were also evident during the pre-symptomatic period (n = 117 multiple sclerosis; n = 60 controls). Prior to clinical onset of multiple sclerosis, individuals exhibited a higher systemic age gap of 1.0 years (95% CI 0.4 to 1.7; *P* = 0.002; FDR= 0.02) and a higher brain age gap of 2.4 years (95% CI 1.2 to 3.7; *P* < 0.001; FDR = 0.002). Higher proteomic age gap was also observed for the liver (β = 2.0 years; 95% CI 0.57 to 3.50; *P* = 0.007; FDR = 0.04) and artery (β = 0.9 years; 95% CI 0.04 to 1.80; *P* = 0.04; FDR = 0.15) at this timepoint. In contrast, there was no evidence of increased immune age gap in either the pre-symptomatic period (β = 0.5 years; 95% CI −0.5 to 1.4; *P* = 0.40; FDR = 0.62) or the post-onset period (median 2 years after onset; β = 0.07 years; 95% CI −0.8 to 1.0; *P* = 0.90; FDR = 0.96) (Figure 3). No significant differences were observed for the remaining organ-specific proteomic age gap measures in either the pre-symptomatic or post-onset samples.

In the JHU cohort, male sex was associated with higher proteomic age gaps in the brain (β = 1.0 years; 95% CI 0.1 to 1.8; *P* = 0.02; FDR = 0.09), muscle (β = 1.0 years; 95% CI 0.1 to 1.9; P = 0.03; FDR = 0.09), and kidney (β = 0.5 years; 95% CI 0.07 to 1.0; *P* = 0.02; FDR = 0.09), independent of age, race, and multiple sclerosis subtype. Male sex was also associated with a lower artery (β = −1.5 years; 95% CI −2.4 to −0.6; *P* < 0.001; FDR = 0.01) and organismal age gap (β = −0.8 years; 95% CI −1.6 to −0.01; *P* = 0.05; FDR = 0.12). No significant differences were observed for systemic (β = −0.6 years; 95% CI −1.4 to 0.1; *P* = 0.10; FDR = 0.18), immune (β = 0.5 years; 95% CI −0.4 to 1.4; *P* = 0.30; FDR = 0.48), and other organ-specific age gaps.

In the DoDSR cohort, age-adjusted analyses demonstrated higher proteomic age gaps in males for systemic (β = 1.9 years; 95% CI 1.0 to 2.7; *P* < 0.001; FDR <0.001), pancreas (β = 1.9 years; 95% CI 0.8 to 3.1; *P* = 0.001; FDR = 0.006), muscle (β = 1.4 years; 95% CI 0.4 to 2.4; *P* = 0.005; FDR = 0.02), kidney (β = 0.7 years; 95% CI 0.3 to 1.1; *P* < 0.001; FDR = 0.004), and intestine (β = 1.7 years; 95% CI 0.2 to 3.2; *P* = 0.03; FDR = 0.08). Associations for brain (β = 0.8 years; 95% CI −0.7 to 2.4; *P* = 0.28; FDR = 0.37), immune (β = 1.0 years; 95% CI −0.1 to 2.2; *P* = 0.08; FDR = 0.15), and other organ-specific age were not statistically significant.

In the JHU cohort, compared to relapsing remitting, progressive multiple sclerosis was associated with a higher systemic age gap (β = 1.2 years; 95% CI 0.4 to 1.9; *P* = 0.003; FDR = 0.02), organismal age gap (β = 1.1 years; 95% CI 0.3 to1.9; *P* = 0.009; FDR = 0.04), and muscle age gap (β = 1.8 years; 95% CI 0.9 to 2.7; *P* = 0.0001; FDR = 0.002). No significant differences were observed for brain, (β = 0.4 years; 95% CI −0.5 to 1.2; *P* = 0.40; FDR = 0.66), immune (β = 0.5 years; 95% CI −0.4 to 1.4; *P* = 0.29; FDR = 0.63) and other organ-specific age gaps.

In the JHU cohort, DMT class was not associated with either systemic proteomic age gap (*P* = 0.49) or immune proteomic age gap (*P* = 0.12) among individuals receiving treatment, and there was no evidence of effect modification by race for the systemic age gap (*P* = 0.50).

No significant association was observed between systemic proteomic age gap (*P* = 0.11) or brain proteomic age gap (*P* = 0.90) and time to first clinical symptom onset in the DoDSR cohort.

### Association of proteomic age gap with multiple sclerosis outcome measures

Significant associations between systemic and organ-specific proteomic age gaps and multiple sclerosis clinical and imaging outcomes were observed after correction for multiple testing (FDR < 0.05) (Figure 4).

**Figure 4.**
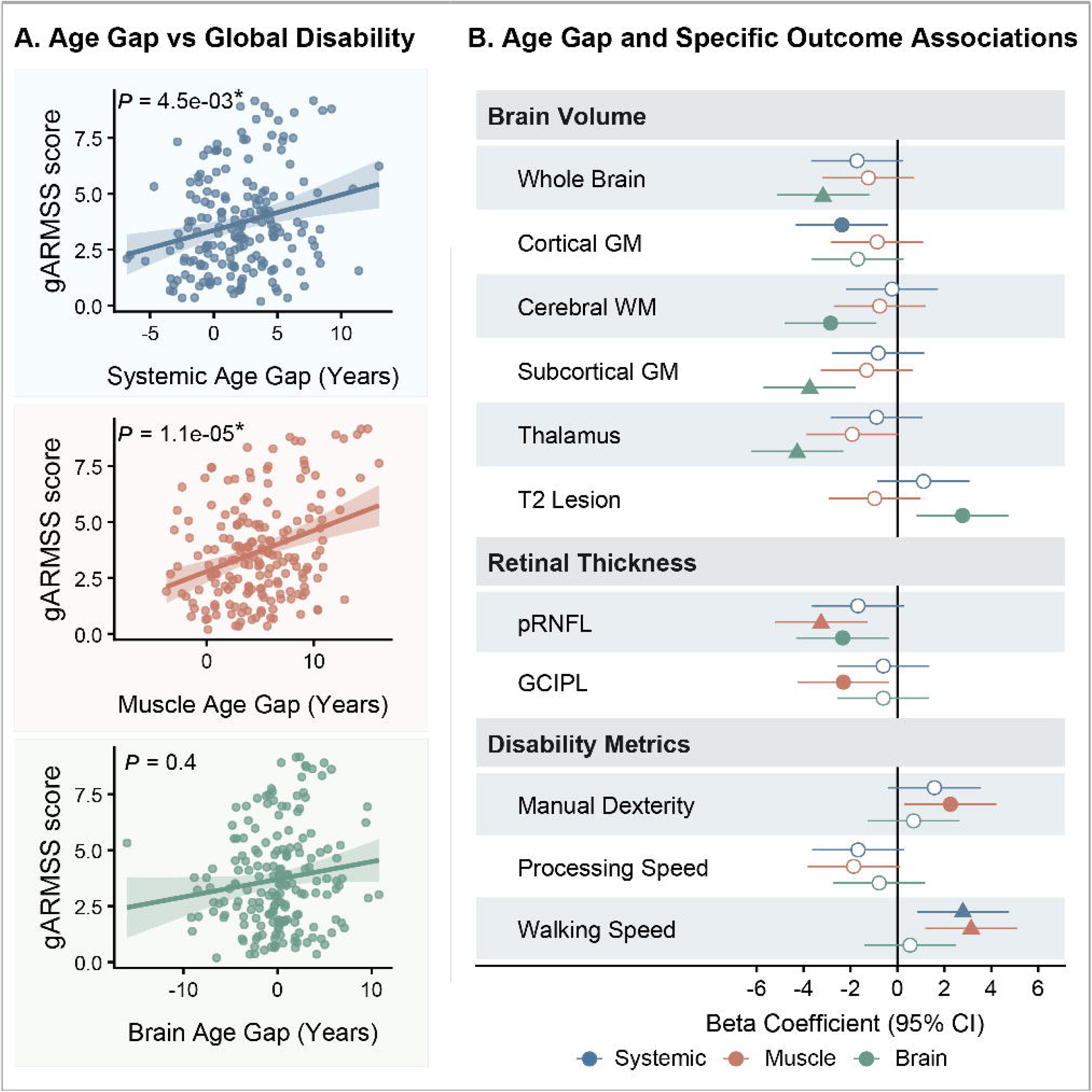
Associations of proteomic age gaps with clinical disability and imaging outcomes in the Johns Hopkins cohort. **(A)** Associations between systemic, muscle, and brain proteomic age gaps and global Age-Related Multiple Sclerosis Severity Score (gARMSS). Higher systemic and muscle proteomic age gaps were associated with greater gARMSS. P values are derived from linear regression models adjusted for chronological age, sex, and race. Asterisks (*) denote associations that remained significant after false discovery rate (FDR) correction (FDR-adjusted *P* < 0.05). **(B)** Associations between proteomic age gaps and imaging and clinical outcomes. Higher systemic proteomic age gap was associated with slower walking speed, and lower cortical grey matter volume. Higher muscle proteomic age gap was associated with poorer manual dexterity, reduced walking speed, and lower retinal thicknesses. Higher brain proteomic age gap was associated with lower whole-brain volume, cerebral white matter, subcortical grey matter, and thalamic volume, as well as greater T2 lesion volume and lower retinal thickness. Points represent beta coefficients with 95% confidence intervals from multivariable linear regression models adjusted for chronological age, sex, and race. Filled circles denote nominally significant associations (*P* < 0.05), and triangles denote associations that remained significant after FDR correction. Abbreviations: gARMSS, Age-Related Multiple Sclerosis Severity Score; pRNFL, peripapillary retinal nerve fiber layer thickness; GCIPL, ganglion cell–inner plexiform layer thickness.

#### Clinical disability and functional performance

Higher systemic age gap was associated with greater disability severity, as measured by gARMSS (β = 0.14, 95% CI 0.05 to 0.24, *P* = 0.005, FDR = 0.03) and poorer walking performance measured by the walking speed test (β = 0.02, 95% CI 0.01 to 0.03, *P* = 0.006, FDR = 0.04). Proteins contributing most strongly to the systemic aging signal included GDF15, IGDCC4, ENPP5, MSR1, CA3, CLEC3B, CR2, TREM1, WFDC2, and CTSV (Figure 5, Supplementary table 1-2). Pathway enrichment analysis of these proteins highlighted processes related to mitochondrial organization, small molecule metabolic processes, and intrinsic apoptotic signaling pathways, along with cellular components related to mitochondria and synapse-associated extracellular matrix, and molecular functions such as oxidoreductase activity and cofactor binding (Figure 5).

**Figure 5.**
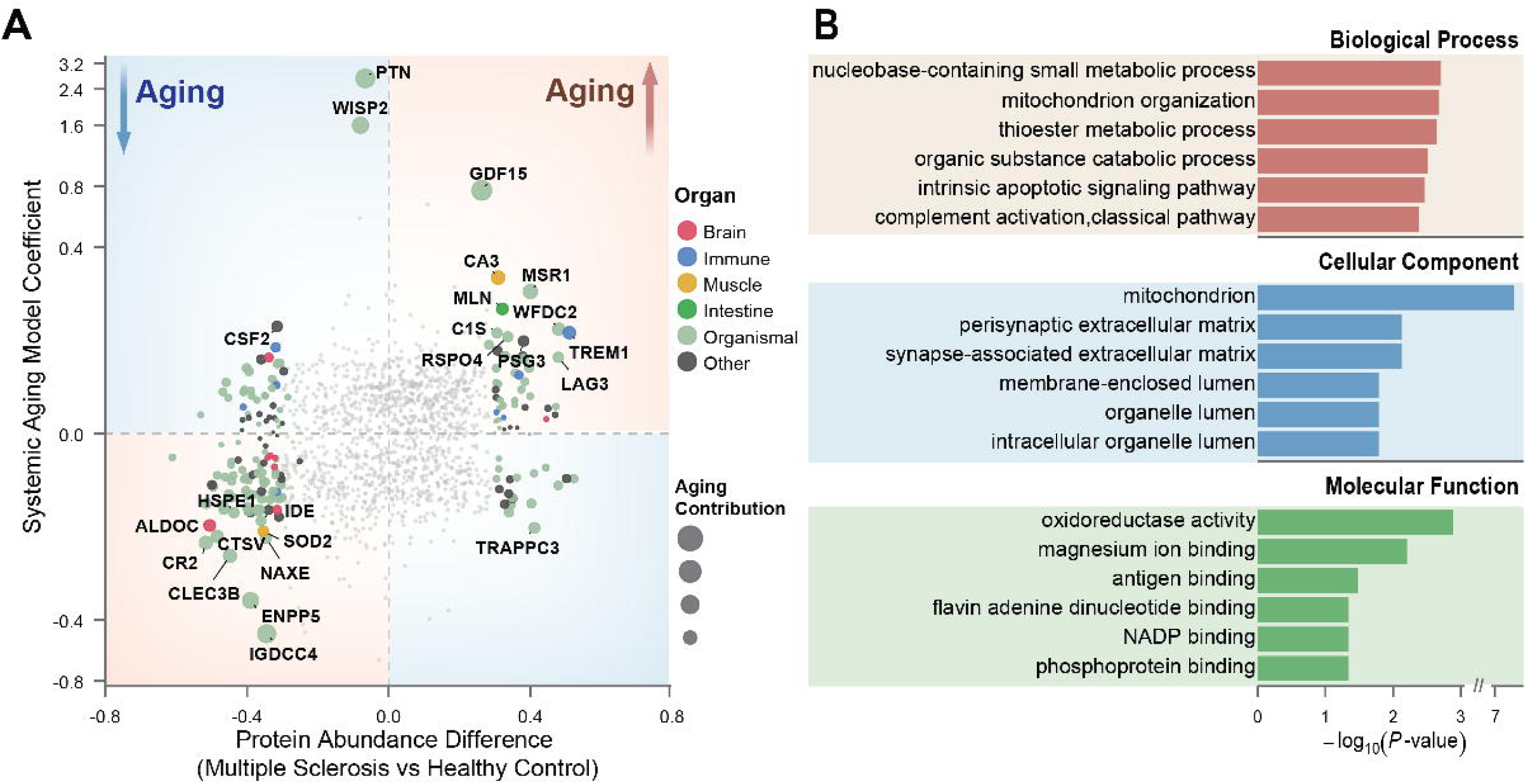
Proteins contributing to systemic aging in multiple sclerosis and pathway enrichment underlying proteomic age acceleration in multiple sclerosis. **(A)** Top proteins contributing to systemic proteomic aging in people with multiple sclerosis relative to healthy controls. Protein contribution was defined as the product of normalized protein abundance and the corresponding coefficient from the systemic proteomic aging model, representing the protein’s weighted contribution to predicted biological age. Dot size represents the absolute value of the estimated regression coefficient from multivariable linear regression models fitted separately for each protein, with protein contribution as the dependent variable and multiple sclerosis status as the primary independent variable, adjusted for chronological age, sex, and race. Red background shading indicates proteins contributing to accelerated aging, whereas blue background shading indicates proteins contributing to decelerated aging. Dot colors indicate organ systems. **(B)** Gene Ontology pathway enrichment analysis of proteins significantly (*P* < 0.05) contributing to accelerated aging, highlighting enriched biological processes, cellular components, and molecular functions.

Higher muscle age gap was associated with greater gARMSS (β = 0.17, 95% CI 0.10 to 0.24, *P* < 0.001, FDR < 0.001), reduced walking speed (β = 0.02, 95% CI 0.01 to 0.03, *P* = 0.002, FDR = 0.02), and poorer manual dexterity measured by the nine-hole peg test (β = 0.28, 95% CI 0.04 to 0.52, *P* = 0.03, FDR = 0.30). The muscle age acceleration was predominantly driven by CA3, SOD2, RAD23A, and TTN (Supplementary table 1-2).

#### Volumetric brain MRI measures

Proteomic aging was also associated with structural MRI measures. Higher brain age gap was associated with lower whole-brain volume (β = −1.5×10□³, 95% CI −2.5×10□³ to −5.8×10□□, *P* = 0.002, FDR = 0.02), cerebral white matter volume (β = −7.7×10□□, 95% CI −1.3×10□³ to −2.4×10□□, *P* = 0.005, FDR = 0.06), subcortical gray matter volume (β = −1.5×10□□, 95% CI −2.3×10□□to −7.2×10□□, *P* <0.001, FDR = 0.03), and thalamic volume (β = −5.9×10□□, 95% CI −8.6×10□□to −3.2×10□□, *P* <0.001, FDR <0.001).

Higher brain age gap was also associated with greater T2 lesion volume (β = 2.38 × 10□□; 95% CI 6.9 × 10□□to 4.1 × 10□□; *P* = 0.006, FDR= 0.1). Additionally, higher systemic age gap was associated with lower cortical gray matter volume (β = −7.1×10□□, 95% CI −1.3×10□³ to −1.2×10□□□, *P* = 0.02, FDR= 0.10). Proteins driving the brain aging included several proteins, notably ALDOC, LANCL1, AGAP2, GPC5, PAK5, OMG, TNR, KLC1, BCAN, and GAP43 (Supplementary table 1-2).

#### Retinal thicknesses

Higher brain proteomic age gap was associated with lower pRNFL thickness (β = −0.21, 95% CI −0.39 to −0.03, *P* = 0.02, FDR= 0.10). Similarly, higher muscle age gap was associated with lower pRNFL thickness (β = −0.26, 95% CI −0.41 to −0.10, *P* = 0.001, FDR = 0.02) and GCIPL thickness (β = −0.35; 95% CI −0.65 to −0.05; *P* = 0.02; FDR = 0.30).

Overall, systemic proteomic age gap and muscle age gap were primarily linked to global disability measures and functional disability, while brain-specific aging was more so associated with volumetric MRI and retinal OCT measures (Figure 4). In contrast, immune proteomic age gap was not associated with clinical disability or imaging outcomes in this cohort. Detailed results of organ-specific associations and a complete ranked list of protein contributors for each aging clock are provided in Supplementary Tables 1-2.

## Discussion

Using two independent cohorts spanning distinct proteomic platforms, we demonstrate that systemic and organ-specific proteomic aging are accelerated in multiple sclerosis compared with age-matched controls. Notably, systemic, brain, and liver proteomic age acceleration were detectable several years before clinical onset, extending prior observations of biological aging in multiple sclerosis by clarifying the temporal relationship between biological aging and multiple sclerosis^4–7^. These findings suggest that dysregulated aging biology may represent an early component of multiple sclerosis pathogenesis rather than solely a consequence of established disease. In contrast, immune proteomic age acceleration was not evident during the pre-symptomatic or early post-onset phases, despite the autoimmune nature of the disease. Notably, in individuals with established multiple sclerosis, immune aging was relevant suggesting that immune aging may develop later in the disease course. Importantly, higher proteomic age gaps were associated with greater disability, poorer functional performance, as well as reduced MRI-derived brain volumes and retinal thicknesses. These findings support the clinical relevance of proteomic aging signatures and link them to pathological processes in multiple sclerosis.

A major finding of this study is the detection of proteomic age acceleration in pre-symptomatic individuals who later developed multiple sclerosis, indicating that biological aging processes are altered before symptomatic neuroinflammation becomes clinically apparent. This extends prior observations by showing that accelerated biological aging is not merely a consequence of long-standing disease or exposure to disease-modifying therapies but may emerge before clinical onset. This result is consistent with growing evidence that multiple sclerosis is preceded by a prolonged prodromal phase characterized by measurable molecular and physiological changes, including alterations in serum neurofilament light chain, autoantibody signatures, and healthcare utilization patterns, several years before the first clinical demyelinating event^15,31,32^. Consistent with prior studies, our findings suggest that this prodromal phase may include broader biological changes that extend beyond canonical neuroinflammatory markers. Notably, brain-associated proteins contributed prominently to the pre-symptomatic aging signal, aligning with evidence that neuroaxonal injury and blood–brain barrier disruption are among the earliest detectable abnormalities in multiple sclerosis pathogenesis^15,31,33^.

While the pre-symptomatic detection of proteomic age acceleration raises important questions regarding causality; the design of the DoDSR cohort does not allow these questions to be fully resolved. One possibility is that accelerated biological aging represents a pre-existing state that increases susceptibility to multiple sclerosis. An alternative explanation is that subclinical pathological processes, including low-grade neuroinflammation, neuroaxonal injury, and altered neurovascular integrity, drive premature aging signals in the circulating proteome. Several observations favor the latter interpretation. Brain-specific proteomic aging was more pronounced than systemic aging during the pre-symptomatic phase; age acceleration was not associated with proximity to symptom onset; and it was not associated with DMT class. These findings argue against a simple model in which aging signals merely accumulate with overt disease burden and instead suggest that aging-related processes may already be embedded within the earliest stages of multiple sclerosis biology. This interpretation is consistent with prior reports of accelerated biological aging markers, including telomere shortening and epigenetic age acceleration, in treatment-naïve and pediatric-onset multiple sclerosis ^5,34^. Taken together, the data support the idea that aging-related molecular processes may precede and interact with disease pathophysiology, rather than arising solely as downstream consequences of established neuroinflammation or accumulated disability.

Our findings also refine current understanding of immune aging in multiple sclerosis. Consistent with previous studies, we observed immune proteomic age acceleration in individuals with established disease^34–37^. However, immune proteomic aging was not detected during the pre-symptomatic or early post-onset phases, in contrast to the brain and systemic proteomic aging signals that were already evident at these stages. The divergent immune aging findings across the two cohorts warrant careful interpretation. Immune age acceleration was evident in the JHU cohort, composed predominantly of older individuals with established multiple sclerosis, but not in the DoDSR cohort, which comprised younger military personnel sampled during the pre-symptomatic and early disease phases. This discrepancy may reflect true stage-dependent differences in immune aging, or differences in protein coverage between proteomic aging models, as well as potential differences in sample stability related to longer-term storage in the DoDSR cohort, with some proteins driving immune aging being less stable. Immunosenescence is a well-established feature of multiple sclerosis and includes expansion of senescent T-cell populations, depletion of naïve T cells, and chronic low-grade inflammation^33,38,39^. Our data suggest, however, that these immune aging features may become more prominent with disease duration and chronological age rather than being a hallmark of pre-symptomatic disease.

Although our data do not directly establish mechanism, pathway enrichment points to mitochondrial dysfunction as a key mechanism underlying proteomic aging in multiple sclerosis. Mitochondrial damage accumulates with age and is amplified by chronic inflammation, resulting in energy failure, excess reactive oxygen species generation, and activation of cell death pathways^40–42^. In multiple sclerosis, mitochondrial dysfunction has been implicated in axonal degeneration, oligodendrocyte injury, and impaired remyelination^33^. More broadly, interactions among chronic neuroinflammation, immunosenescence, mitochondrial dysfunction, and systemic metabolic stress may create a self-reinforcing network that amplifies biological aging across tissues.

Muscle proteomic aging emerged as a particularly strong correlate of clinical impairment in the JHU cohort. Muscle age gap was associated with global disability, motor dysfunction, and retinal thinning, suggesting that muscle aging may contribute to disability accumulation through both direct effects on motor performance and shared biological pathways affecting CNS integrity. Among the proteins driving this signal, CA3 (carbonic anhydrase 3) is of particular interest. CA3 is highly enriched in slow-twitch skeletal muscle fibers and plays key roles in pH regulation, antioxidant defense, and fatigue resistance^43–45^. Increased CA3 expression has been reported in aging skeletal muscle and has been linked to fiber-type shifts toward slower, more oxidative phenotypes^44,46^. In aging and neurodegenerative disorders, the muscle–brain axis has emerged as an important mediator of motor and cognitive function through myokine signaling, mitochondrial function, and shared inflammatory pathways^47,48^. Our results suggest that related mechanisms may be relevant in multiple sclerosis, potentially involving impaired muscle-derived neurotrophic support, and convergent mitochondrial stress in muscle and CNS tissues^41,42,47,48^. Further investigation of muscle mitochondrial function, myokine secretion, and muscle-derived trophic signaling in multiple sclerosis may therefore reveal new therapeutic targets and inform rehabilitation strategies. Importantly, the observed muscle age acceleration and its association with clinical outcomes may reflect, at least in part, a consequence of disease-related disability. Reduced physical activity, deconditioning, and muscle atrophy associated with higher disability levels in multiple sclerosis may contribute to these signals. Given the cross-sectional design, causality cannot be determined, and longitudinal studies are required to clarify these relationships.

These observations suggest several potential clinical implications. Proteomic aging clocks may serve as biomarkers of disease severity and as prognostic tools to identify individuals at higher risk of disability progression, with potential applications in guiding treatment strategies and enriching clinical trial populations. The pre-symptomatic detection of proteomic age acceleration further raises the possibility that these biomarkers could support early disease detection in high-risk populations, such as individuals with radiologically isolated syndrome or family members of people with multiple sclerosis. In addition, the proteins and pathways contributing to age acceleration may represent potential therapeutic, raising the possibility that interventions targeting biological aging could modify disease progression^49–51^. Candidate strategies include senolytics to eliminate senescent cells, senomorphics to suppress the senescence-associated secretory phenotype, mitochondrial-targeted therapies, and anti-inflammatory approaches directed at specific aging-related pathways. Lifestyle interventions, including exercise and dietary modification, may also be relevant given their established effects on healthy aging and systemic metabolic resilience, as well as emerging evidence of reducing metabolomic aging in multiple sclerosis^49–53^. Although these possibilities remain speculative, our findings highlight the potential value of further investigating aging biology not only as a contributor to multiple sclerosis progression but also as a potential avenue for therapeutic intervention.

This study has several important strengths. These include the use of two independent cohorts with distinct demographic characteristics and proteomic platforms, the inclusion of pre-symptomatic samples providing temporal insight, the availability of detailed clinical and imaging phenotypes, and the application of validated proteomic aging clocks derived from large population-based datasets^54–57^. The convergence of key findings across cohorts and platforms strengthens confidence that observed signals are biologically meaningful rather than being platform-specific artifacts.

Certain limitations should also be acknowledged. Limited longitudinal data in both cohorts restricted the analyses largely to cross-sectional comparisons, limiting causal inference regarding the relationships between proteomic aging, disease onset, and progression. Longitudinal studies integrating proteomic, imaging, and clinical trajectories will be necessary to define the temporal dynamics of biological aging in multiple sclerosis and determine whether these processes are modifiable by treatment. The DoDSR cohort consisted primarily of young, predominantly male military personnel, which may limit generalizability to the broader multiple sclerosis population. In addition, differences in proteomic aging models between cohorts, including variations in protein coverage, platforms, and model derivation, may contribute to differences in observed aging signals and limit direct comparability between cohorts. Differences in sample collection, processing, and storage conditions between cohorts may also influence measured protein levels and derived aging estimates across cohorts. Although we identified proteins contributing to the aging signals, further validation will be required to determine whether these proteins are causal drivers of multiple sclerosis pathogenesis or downstream markers of disease-related processes.

In conclusion, this study demonstrates that systemic and organ-specific proteomic age acceleration is present in multiple sclerosis and can be detected years before clinical symptom onset. The identification of these proteomic aging signatures in the pre-symptomatic phase suggests that accelerated biological aging may represent an early component of multiple sclerosis pathophysiology rather than simply a consequence of established disease. Brain-specific proteomic aging was particularly pronounced and associated with structural brain measures, whereas muscle-specific aging was strongly linked to disability and functional impairment. Together, these findings position proteomic age as a promising biological readout of disease processes and a candidate diagnostic and prognostic biomarker. More broadly, they support the concept that aging biology is integral to multiple sclerosis pathogenesis and may offer a new framework for risk stratification, mechanistic discovery, and therapeutic intervention.

## Data availability

The study’s anonymized data will be available from the corresponding author upon reasonable request. The complete proteomic dataset generated from the DODSR participants and related demographics, and clinical data are available via Dryad at https://doi.org/10.5061/dryad.fttdz093t11.

